# Learning as We Go: An Examination of the Statistical Accuracy of COVID19 Daily Death Count Predictions

**DOI:** 10.1101/2020.04.11.20062257

**Authors:** Roman Marchant, Noelle I. Samia, Ori Rosen, Martin A. Tanner, Sally Cripps

## Abstract

A recent model developed at the Institute for Health Metrics and Evaluation (IHME) provides forecasts for ventilator use and hospital beds required for the care of COVID19 patients on a state-by-state basis throughout the United States over the period March 2020 through August 2020 (See the related website https://covid19.healthdata.org/projections for interactive data visualizations). In addition, the manuscript and associated website provide projections of deaths per day and total deaths throughout this period for the entire US, as well as for the District of Columbia. This research has received extensive attention in social media, as well as in the mass media. Moreover, this work has influenced policy makers at the highest levels of the United States government, having been mentioned at White House Press conferences, including March 31, 2020.

In this paper, we evaluate the predictive validity of model forecasts for COVID19 outcomes as data become sequentially available, using the IHME prediction of daily deaths. We have found that the predictions for daily number of deaths provided by the IHME model have been highly inaccurate. The model has been found to perform poorly even when attempting to predict the number of next day deaths. In particular, the true number of next day deaths has been outside the IHME prediction intervals as much as 70% of the time.

## 1 Introduction

A recent model developed at the Institute for Health Metrics and Evaluation (IHME) provides forecasts for ventilator use and hospital beds required for the care of COVID19 patients on a state-by-state basis throughout the United States over the period March 2020 through August 2020 [4] (See also the related website https://covid19.healthdata.org/projections for interactive data visualizations). In addition, the manuscript and associated website provide projections of deaths per day and total deaths throughout this period for the entire US, as well as for the District of Columbia. This research has received extensive attention in social media, as well as in the mass media [2, 3]. Moreover, this work has influenced policy makers at the highest levels of the United States government, having been mentioned at White House Press conferences, including March 31, 2020 [2].

Our goal in this report is to provide a framework for evaluating the predictive validity of model forecasts for COVID19 outcomes as data become sequentially available, using the IHME prediction of daily deaths as an example. Given our goal is to provide an evaluation framework, we treat the IHME model as a “black box” and examine the projected numbers of deaths per day in light of the ground truth to help begin to understand the predictive accuracy of the model. We do not provide a critique of the assumptions made by the IHME model, nor do we suggest any possible modifications to the IHME approach. Moreover, our analysis should not be misconstrued as an investigation of mitigation measures such as social distancing. As of April 5 2020, IHME released a new version of their model and we will examine the associated predictions in the coming days as more data become available.

## 2 Method

Our report examines the quality of the IHME deaths per day predictions for the period March 29–April 2, 2020. For this analysis we use the actual deaths attributed to COVID19 on March 30 and March 31 as our ground truth. Our source for these data is the number of deaths reported by Johns Hopkins University ([1]).

Each day the IHME model computes a daily prediction and a 95% posterior interval (PI) for COVID19 deaths, four months into the future for each state. Thus, on March 29 there is a prediction and corresponding PI for March 30 and March 31, while on March 30 there is a prediction and corresponding PI for March 31. We call the prediction for a day made on the previous day a “1-step-ahead” prediction. Similarly, a prediction for a day made two days in advance is referred to as a “2-step-ahead” prediction, while a prediction for a day made *k* days in advance is called a “k-step-ahead” prediction.

Using these definitions, for each location we have a 1-step-ahead prediction for March 30, made on March 29; a 2-step-ahead and a 1-step-ahead prediction for March 31, made on March 29 and March 30, respectively. For April 1 we have a 3-step, a 2-step and a 1-step-ahead prediction, made on March 29, 30 and 31, respectively, etc.

## 3 Results

Figure 1 graphically represents the discrepancy between the actual number of deaths and the 95% PIs for deaths, by state for the dates March 30 through to April 2. The color in these figures shows whether the actual death counts were less than the lower limit of the 95% PI (blue), or within the 95% PI (white), or above the upper limit of the 95% PI (red). The depth of the red/blue color denotes the number of actual death counts above/below the PI. A deep red signifies that a large number of deaths were above the upper limit of the 95% PI, while a light red indicates a smaller number. Similarly, a deep blue signifies that a large number of deaths were below the lower limit of the 95% PI, while a light blue indicates a smaller number.

**Figure 1:**
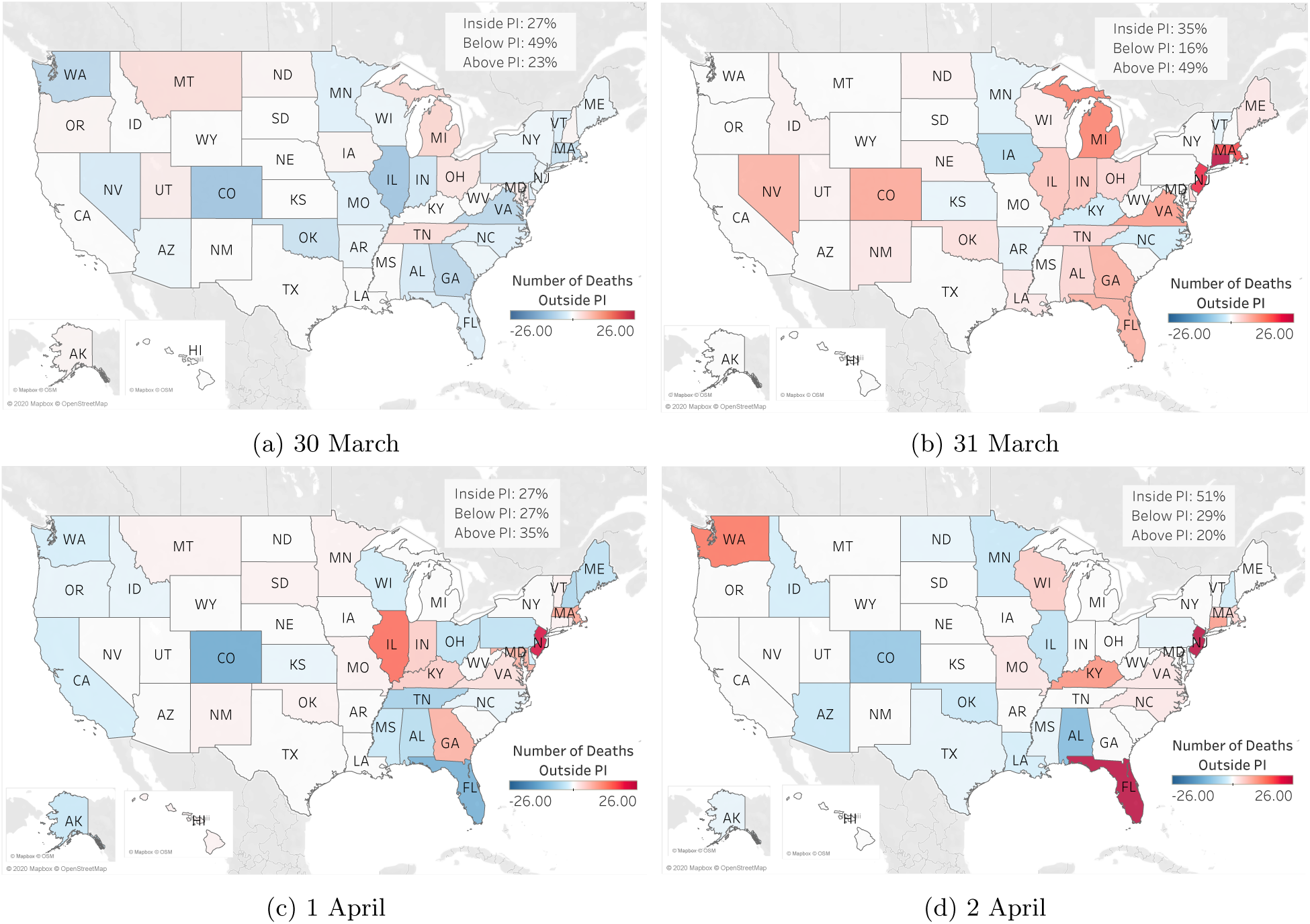
Discrepancy between actual death counts and one-step-ahead PIs for specific dates (see sub-figures). The color shows whether the actual death counts were less than the lower limit of the 95% PI (blue), within the 95% PI (white), or above the upper limit of the 95% PI (red). The depth of the red/blue color denotes how many actual deaths were above/below the 95% PI.

These figures show that for March 30 only 27% of states had an actual number of deaths lying in the 95% PI for the 1-step-ahead forecast. The corresponding percentages for March 31, April 1 and April 2, are 35%, 27% and 51%, respectively. Therefore percentage of states with actual number of deaths lying outside this interval is 73%, 65%, 73% and 49% for March 30, March 31, April 1 and April 2, respectively. We note that we would expect only 5% of observed death counts to lie outside the 95% PI.

For a given day the model is also biased, although the direction of the bias is not constant across days. For the 1-step-ahead prediction for March 30th, 49% of all locations were over-predicted, that is 49% of all locations had a death count which was below the 95% PI lower limit, while 23% were under-predicted. For March 31st the reverse was true; only 16% of locations had actual death counts below the 95% PI lower limit while 49% had actual death counts above the 95% PI upper limit. This can be clearly seen from Figures 1a and 1b which are predominantly blue, and red, respectively. These figures are summarized in Table 1.

**Table 1:** Percentage of locations with actual death counts inside the 95% PI, as a function of the number of forecast periods. The values in parentheses indicate the percentage of locations that were (below,above) the limits of the 95% PI.

|  | 1-step | 2-step | 3-step | 4-step |
| --- | --- | --- | --- | --- |
| March 30 | 27 (49,23) |  |  |  |
| March 31 | 35 (16,49) | 45 (14,41) |  |  |
| April 1 | 27 (27,35) | 39 (29,31) | 43 (43,23) |  |
| April 2 | 51 (29,20) | 51 (25,23) | 45 (25,28) | 47 (25,28) |

Table 1 also shows that the accuracy of predictions does not improve as the forecast horizon decreases, as one would expect. For March 31 and April 1 the forecast accuracy, as measured by the percentage of states whose actual death count lies within the 95% PI, decreases as the forecast horizon decreases. For March 31 the 2-step ahead prediction is better than the 1-step ahead prediction, while for April 1 the 3-step is better than the 2-step, which in turn is better than the 1-step. However April 2 shows that accuracy slightly improves between the 3-step and the 2-step.

To investigate the relationship between the 2-step-ahead and the 1-step-ahead prediction errors by state, Figure 2 shows the March 31 1-step-ahead prediction errors, made on March 30, on the y-axis, versus the March 31 2-step-ahead prediction errors, made on March 29, on the x-axis. The colors in the graph correspond to different subsets of the data; red corresponds to those locations where the actual number of deaths was above the 1-step-ahead 95% PI upper limit, blue corresponds to those locations where the actual number of deaths was below the 1-step-ahead 95% PI lower limit, while grey corresponds to those locations where the actual number of deaths was within the 1-step-ahead 95% PI. This graph shows a very strong linear association between the predicted errors for the red locations (*R*^2^=96% n=25). This suggests that the additional information contained in the March 30 data did little to improve the prediction for those locations where the actual death count was much higher than the predicted number of deaths. The number of observations in the other two subsets of data was insufficient to draw any firm conclusions.

**Figure 2:**
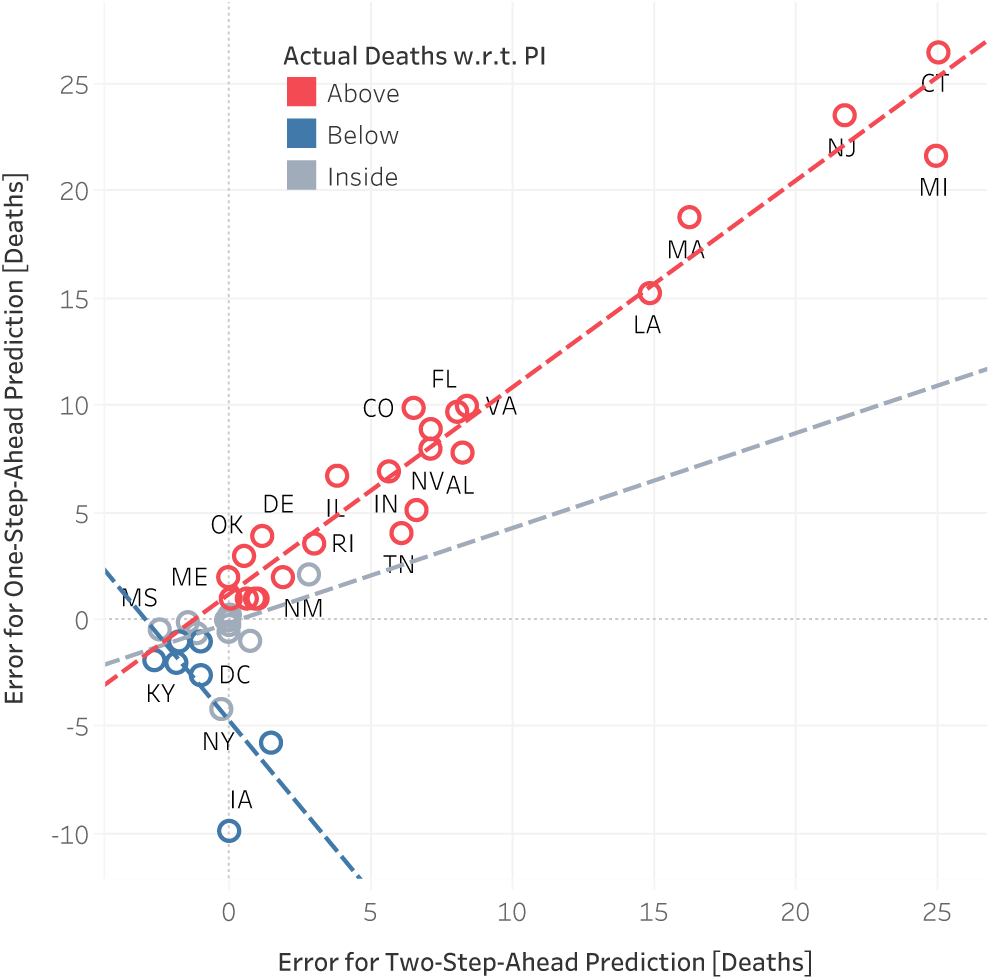
Actual minus predicted values of the 1-step ahead prediction for March 31, (y-axis) vs actual minus predicted value of the 2-step ahead prediction for March 31, (x-axis). The colors in the graph correspond to different subsets of the data; red corresponds to those locations where the actual number of deaths was above the 1-step-ahead 95% PI upper limit, blue corresponds to those locations where the actual number of deaths was below the 1-step-ahead 95% PI lower limit, while grey corresponds to those locations where the actual number of deaths was within the the 1-step-ahead 95% PI.

## 4 Discussion

Our results suggest that the IHME model substantially underestimates the uncertainty associated with COVID19 death count predictions. We would expect to see approximately 5% of the observed number of deaths to fall outside the 95% prediction intervals. In reality, we found that the observed percentage of death counts that lie outside the 95% PI to be in the range 49% - 73%, which is more than an order of magnitude above the expected percentage. Moreover, we would expect to see 2.5% of the observed death counts fall above and below the PI. In practice, the observed percentages were asymmetric, with the direction of the bias fluctuating across days.

We note that there are two ways in which the accuracy of the model, as measured by the percentage of states with death counts which fall within the 95% PI, can improve. Either the estimated uncertainty increases and therefore the prediction intervals become much wider, or the point of the estimated expected value improves. The latter is preferable but much harder to achieve in practice. The former can potentially lead to prediction intervals that are too wide to be useful to drive the development of health, social, and economic policies.

In addition, the performance accuracy of the model does not improve as the forecast horizon decreases. In fact, Table 1 indicates that the reverse is generally true. Interestingly, the model’s prediction for the state of New York is consistently accurate, while the model’s prediction of the neighboring state of New Jersey, which is part of the New York metropolitan area, is not consistently accurate.

We would like to thank the authors of IHME for making their predictions and data publicly available. We agree with the statement on their website http://www.healthdata.org/covid/updates *Having more timely, high-quality data is vital for all modeling endeavors, but its importance is dramatically higher when trying to quantify in real time how a new disease can affect lives*. Without access to the data and predictions this analysis would not have been possible. We look forward to evaluating the performance of the newer versions of the IHME model.

## Data Availability

Data are available on the IHME website below:
https://covid19.healthdata.org/projections
Citation: Christopher JL Murray. Forecasting covid-19 impact on hospital bed-days, icu-days, ventilator-days and deaths by us state in the next 4 months. medRxiv, 2020

